# Bivalent mRNA-1273.214 vaccine effectiveness against SARS-CoV-2 omicron XBB* infections

**DOI:** 10.1101/2023.04.15.23288612

**Authors:** Hiam Chemaitelly, Houssein H. Ayoub, Sawsan AlMukdad, Jeremy Samuel Faust, Patrick Tang, Peter Coyle, Hadi M. Yassine, Asmaa A. Al Thani, Hebah A. Al-Khatib, Mohammad R. Hasan, Zaina Al-Kanaani, Einas Al-Kuwari, Andrew Jeremijenko, Anvar H. Kaleeckal, Ali N. Latif, Riyazuddin M. Shaik, Hanan F. Abdul-Rahim, Gheyath K. Nasrallah, Mohamed G. Al-Kuwari, Adeel A. Butt, Hamad E. Al-Romaihi, Mohamed H. Al-Thani, Abdullatif Al-Khal, Roberto Bertollini, Laith J. Abu-Raddad

**Author notes:** Correspondence to Dr. Hiam Chemaitelly, or Professor Laith J. Abu-Raddad,.

## Abstract

Qatar introduced COVID-19 bivalent vaccination for persons ≥12 years old using the 50-μg mRNA-1273.214 vaccine combining SARS-CoV-2 ancestral and omicron BA.1 strains. We estimated effectiveness of this bivalent vaccine against SARS-CoV-2 infection using a matched, retrospective, cohort study. Matched cohorts included 11,482 persons in the bivalent cohort and 56,806 persons in the no-recent-vaccination cohort. During follow-up, 65 infections were recorded in the bivalent cohort and 406 were recorded in the no-recent-vaccination cohort. None progressed to severe, critical, or fatal COVID-19. Cumulative incidence of infection was 0.80% (95% CI: 0.61-1.07%) in the bivalent cohort and 1.00% (95% CI: 0.89-1.11%) in the no-recent- vaccination cohort, 150 days after the start of follow-up. Incidence during follow-up was dominated by omicron XBB* subvariants including XBB, XBB.1, XBB.1.5, XBB.1.9.1, XBB.1.9.2, XBB.1.16, and XBB.2.3. The adjusted hazard ratio comparing incidence of infection in the bivalent cohort to that in the no-recent-vaccination cohort was 0.75 (95% CI: 0.57-0.97). Bivalent vaccine effectiveness against infection was 25.2% (95% CI: 2.6-42.6%). Effectiveness was 21.5% (95% CI: -8.2-43.5%) among persons with no prior infection and 33.3% (95% CI: - 4.6-57.6%) among persons with prior infection. mRNA-1273.214 reduced incidence of SARS-CoV-2 infection, but the protection was modest at only 25%. The modest protection may have risen because of XBB* immune evasion or immune imprinting effects, or combination of both.

---

In October of 2022, Qatar introduced COVID-19 bivalent vaccination for persons ≥12 years old using the 50-μg mRNA-1273.214 vaccine combining SARS-CoV-2 ancestral and omicron BA.1 strains.^1^ We estimated effectiveness of this bivalent vaccine against SARS-CoV-2 infection.

This was done using a matched, retrospective, cohort study to compare infection incidence in the national cohort of persons who received the vaccine (bivalent cohort) to that in the national cohort of Qatar residents who received their last vaccine dose ≥6 months before the start of follow-up (no-recent-vaccination cohort; Section S1 of Supplementary Appendix). The 6-month cut-off was chosen because effectiveness against omicron infection of first-generation vaccines is negligible ≥6 months after the vaccine dose.^2^ Data on SARS-CoV-2 laboratory testing, clinical infection, vaccination, and demographic characteristics were extracted from Qatar’s SARS-CoV- 2 databases.

Incidence of infection was defined as the first SARS-CoV-2 PCR-positive or rapid-antigen-positive test after the start of follow-up, regardless of symptoms. Cohorts were matched exactly by pre-determined factors to balance observed confounders between exposure groups.^3^ Follow-up started 7 days after the person in the bivalent cohort received their vaccine dose. Associations were estimated using Cox proportional-hazards regression models. Hazard ratios were adjusted for the matching factors and testing rate.

Figure S1 shows the study population selection process. Table S1 describes baseline characteristics of full and matched cohorts. Matched cohorts included 11,482 persons in the bivalent cohort and 56,806 persons in the no-recent-vaccination cohort. Median age was 36 years and <5% of study participants were ≥60 years of age. For both cohorts, median duration between last dose, before bivalent dose, and start of follow-up was >1 year.

During follow-up, 65 infections were recorded in the bivalent cohort and 406 were recorded in the no-recent-vaccination cohort (Figure S1). None progressed to severe, critical, or fatal COVID-19. Cumulative incidence of infection was 0.80% (95% CI: 0.61-1.07%) in the bivalent cohort and 1.00% (95% CI: 0.89-1.11%) in the no-recent-vaccination cohort, 150 days after the start of follow-up (Figure 1A). Incidence during follow-up was dominated by omicron XBB, XBB.1, XBB.1.5, XBB.1.9.1, XBB.1.9.2, XBB.1.16, and XBB.2.3.

**Figure 1.**
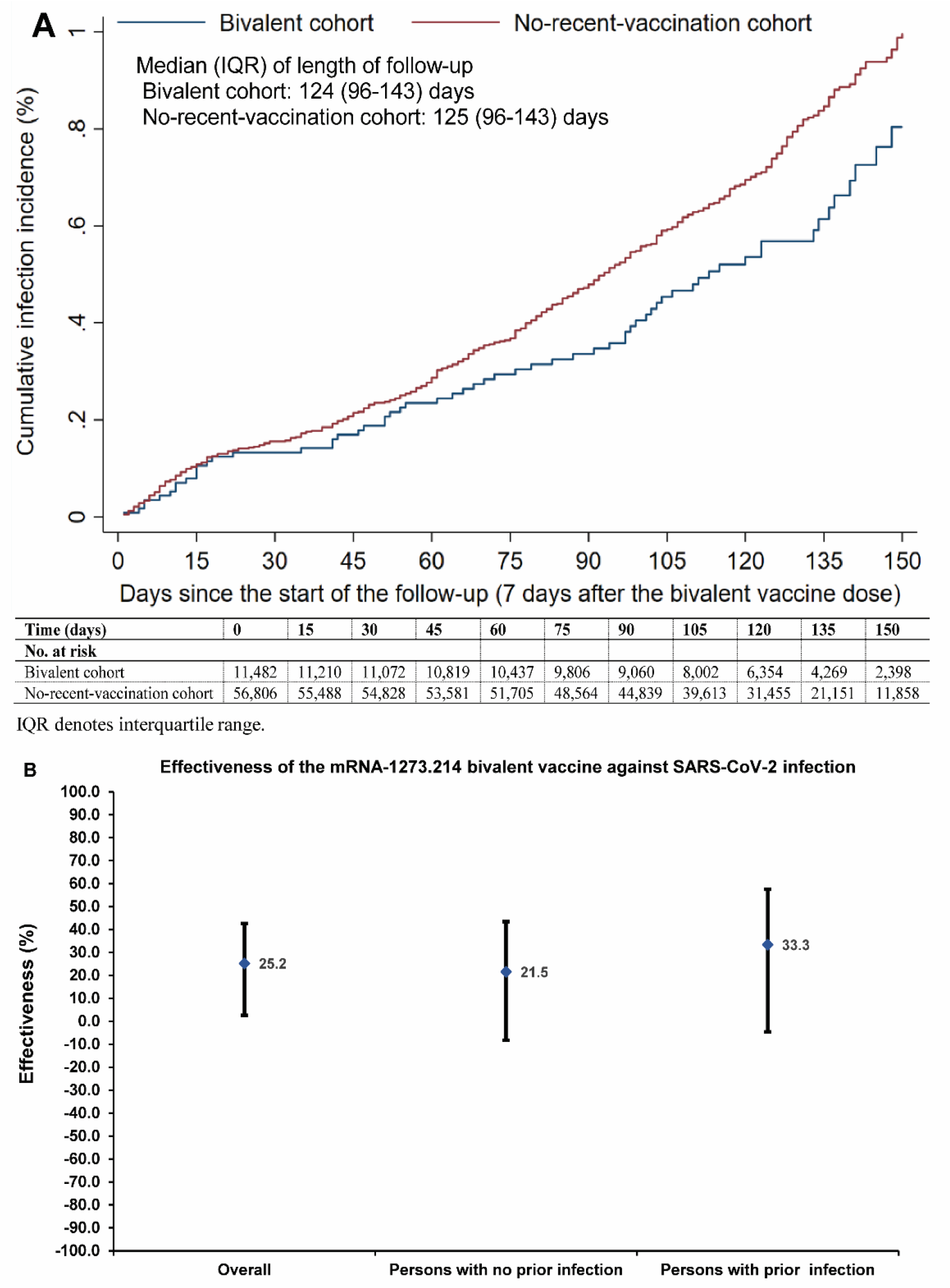
A) Cumulative incidence of SARS-CoV-2 infection in the matched bivalent cohort and the no-recent-vaccination cohort. B) mRNA-1273.214 bivalent vaccine effectiveness against SARS-CoV-2 infection overall and by documented prior infection status. Cohorts were matched exactly one-to-five by sex, 10-year age group, nationality, number of coexisting conditions, and documented prior infection status to balance observed confounders between exposure groups.

The adjusted hazard ratio comparing incidence of infection in the bivalent cohort to that in the no-recent-vaccination cohort was 0.75 (95% CI: 0.57-0.97; Table S2). Bivalent vaccine effectiveness against infection was 25.2% (95% CI: 2.6-42.6%; Figure 1B). Effectiveness was 21.5% (95% CI: -8.2-43.5%) among persons with no prior infection and 33.3% (95% CI: -4.6- 57.6%) among persons with prior infection. In absence of severe COVID-19 cases, effectiveness against severe COVID-19 could not be estimated. Further results and limitations are in Section S2.

mRNA-1273.214 reduced incidence of SARS-CoV-2 infection, but the protection was modest at only 25%, consistent with a modest protection for the bivalent vaccines based on SARS-CoV-2 ancestral and omicron BA.1 or BA.4/BA.5 strains.^4-6^ The modest protection may have risen because of XBB* immune evasion or immune imprinting effects,^2,7,8^ or combination of both.

## Oversight

The institutional review boards at Hamad Medical Corporation and Weill Cornell Medicine– Qatar approved this retrospective study with a waiver of informed consent. The study was reported according to the Strengthening the Reporting of Observational Studies in Epidemiology (STROBE) guidelines (Table S3). The authors vouch for the accuracy and completeness of the data and for the fidelity of the study to the protocol. Data used in this study are the property of the Ministry of Public Health of Qatar and were provided to the researchers through a restricted-access agreement for preservation of confidentiality of patient data. The funders had no role in the study design; the collection, analysis, or interpretation of the data; or the writing of the manuscript.

## Data Availability

The dataset of this study is a property of the Qatar Ministry of Public Health that was provided to the researchers through a restricted-access agreement that prevents sharing the dataset with a third party or publicly. Future access to this dataset can be considered through a direct application for data access to Her Excellency the Minister of Public Health (https://www.moph.gov.qa/english/Pages/default.aspx). Aggregate data are available within the manuscript and its Supplementary information.

## Author contributions

HC co-designed the study, performed the statistical analyses, and co-wrote the first draft of the article. LJA conceived and co-designed the study, led the statistical analyses, and co-wrote the first draft of the article. PVC conducted viral genome sequencing and designed mass PCR testing to allow routine capture of SGTF variants. PT and MRH conducted the multiplex, real-time reverse-transcription PCR variant screening and viral genome sequencing. HY, AAA-T, and HAK conducted viral genome sequencing. All authors contributed to data collection and acquisition, database development, discussion and interpretation of the results, and to the writing of the manuscript. All authors have read and approved the final manuscript.

## Acknowledgements and support

We acknowledge the many dedicated individuals at Hamad Medical Corporation, the Ministry of Public Health, the Primary Health Care Corporation, the Qatar Biobank, Sidra Medicine, and Weill Cornell Medicine – Qatar for their diligent efforts and contributions to make this study possible.

The authors are grateful for support from the Biomedical Research Program and the Biostatistics, Epidemiology, and Biomathematics Research Core, both at Weill Cornell Medicine-Qatar, as well as for support provided by the Ministry of Public Health, Hamad Medical Corporation, and Sidra Medicine. The authors are also grateful for the Qatar Genome Programme and Qatar University Biomedical Research Center for institutional support for the reagents needed for the viral genome sequencing. Statements made herein are solely the responsibility of the authors.

The funders of the study had no role in study design, data collection, data analysis, data interpretation, or writing of the article.

### Supplementary Appendix

#### Section S1. Study population and data sources

Qatar’s national and universal public healthcare system uses the Cerner-system advanced digital health platform to track all electronic health record encounters of each individual in the country, including all citizens and residents registered in the national and universal public healthcare system. Registration in the public healthcare system is mandatory for citizens and residents.

The databases analyzed in this study are data-extract downloads from the Cerner-system that have been implemented on a regular (twice weekly) schedule since onset of the pandemic by the Business Intelligence Unit at Hamad Medical Corporation. Hamad Medical Corporation is the national public healthcare provider in Qatar. At every download all tests, coronavirus disease 2019 (COVID-19) vaccinations, hospitalizations related to COVID-19, and all death records regardless of cause are provided to the authors through .csv files. These databases have been analyzed throughout the pandemic not only for study-related purposes, but also to provide policymakers with summary data and analytics to inform the national response.

Every health encounter in the Cerner-system is linked to a unique individual through the HMC Number that links all records for this individual at the national level. Databases were merged and analyzed using the HMC Number to link all records whether for testing, vaccinations, hospitalizations, and deaths. All deaths in Qatar are tracked by the public healthcare system. All COVID-19-related healthcare was provided only in the public healthcare system. No private entity was permitted to provide COVID-19-related hospitalization. COVID-19 vaccination was also provided only through the public healthcare system. These health records were tracked throughout the COVID-19 pandemic using the Cerner system. This system has been implemented in 2013, before the onset of the pandemic. Therefore, we had all health records related to this study for the full national cohort of Qataris throughout the pandemic. This allowed us to follow each person over time.

Demographic details for every HMC Number (individual) such as sex, age, and nationality are collected upon issuing of the universal health card, based on the Qatar Identity Card, which is a mandatory requirement by the Ministry of Interior to every citizen and resident in the country. Data extraction from the Qatar Identity Card to the digital health platform is performed electronically through scanning techniques.

All severe acute respiratory syndrome coronavirus 2 (SARS-CoV-2) testing in any facility in Qatar is tracked nationally in one database, the national testing database. This database covers all testing in all locations and facilities throughout the country, whether public or private. Every polymerase chain reaction (PCR) test and a proportion of the facility-based rapid antigen tests conducted in Qatar, regardless of location or setting, are classified on the basis of symptoms and the reason for testing (clinical symptoms, contact tracing, surveys or random testing campaigns, individual requests, routine healthcare testing, pre-travel, at port of entry, or other).

Before November 1, 2022, SARS-CoV-2 testing in Qatar was done at a mass scale where about 5% of the population were tested every week.^1,2^ Based on the distribution of the reason for testing up to November 1, 2022, most of the tests in Qatar were conducted for routine reasons, such as being travel-related, and about 75% of cases were diagnosed not because of appearance of symptoms, but because of routine testing.^1,2^

Starting from November 1, 2022, SARS-CoV-2 testing was substantially reduced, but still about 1% of the population are tested every week.^3^ All testing results in the national testing database during follow-up in the present study were factored in the analyses of this study.

The first large omicron wave that peaked in January of 2022 was massive and strained the testing capacity in the country.^1,4-6^ Accordingly, rapid antigen testing was introduced to relieve the pressure on PCR testing. Implementation of this change in testing policy occurred quickly precluding incorporation of reason for testing in a large proportion of the rapid antigen tests for several months. While the reason for testing is available for all PCR tests, it is not available for all rapid antigen tests. Availability of reason for testing for the rapid antigen tests also varied with time.

Rapid antigen test kits are available for purchase in pharmacies in Qatar, but outcome of home-based testing is not reported nor documented in the national databases. Since SARS-CoV-2-test outcomes were linked to specific public health measures, restrictions, and privileges, testing policy and guidelines stress facility-based testing as the core testing mechanism in the population. While facility-based testing is provided free of charge or at low subsidized costs, depending on the reason for testing, home-based rapid antigen testing is de-emphasized and not supported as part of national policy.

Further descriptions of the study population and the national databases were reported previously.^1,2,5,7-10^

#### Section S2. Laboratory methods and variant ascertainment

##### Real-time reverse-transcription polymerase chain reaction testing

Nasopharyngeal and/or oropharyngeal swabs were collected for polymerase chain reaction (PCR) testing and placed in Universal Transport Medium (UTM). Aliquots of UTM were: 1) extracted on KingFisher Flex (Thermo Fisher Scientific, USA), MGISP-960 (MGI, China), or ExiPrep 96 Lite (Bioneer, South Korea) followed by testing with real-time reverse-transcription PCR (RT-qPCR) using TaqPath COVID-19 Combo Kits (Thermo Fisher Scientific, USA) on an ABI 7500 FAST (Thermo Fisher Scientific, USA); 2) tested directly on the Cepheid GeneXpert system using the Xpert Xpress SARS-CoV-2 (Cepheid, USA); or 3) loaded directly into a Roche cobas 6800 system and assayed with the cobas SARS-CoV-2 Test (Roche, Switzerland). The first assay targets the viral S, N, and ORF1ab gene regions. The second targets the viral N and E- gene regions, and the third targets the ORF1ab and E-gene regions.

All PCR testing was conducted at the Hamad Medical Corporation Central Laboratory or Sidra Medicine Laboratory, following standardized protocols.

##### Rapid antigen testing

Severe acute respiratory syndrome coronavirus 2 (SARS-CoV-2) antigen tests were performed on nasopharyngeal swabs using one of the following lateral flow antigen tests: Panbio COVID- 19 Ag Rapid Test Device (Abbott, USA); SARS-CoV-2 Rapid Antigen Test (Roche, Switzerland); Standard Q COVID-19 Antigen Test (SD Biosensor, Korea); or CareStart COVID- 19 Antigen Test (Access Bio, USA). All antigen tests were performed point-of-care according to each manufacturer’s instructions at public or private hospitals and clinics throughout Qatar with prior authorization and training by the Ministry of Public Health (MOPH). Antigen test results were electronically reported to the MOPH in real time using the Antigen Test Management System which is integrated with the national Coronavirus Disease 2019 (COVID-19) database.

##### Classification of infections by variant type

Surveillance for SARS-CoV-2 variants in Qatar is based on viral genome sequencing and multiplex RT-qPCR variant screening^11^ of random positive clinical samples,^2,12-16^ complemented by deep sequencing of wastewater samples.^14,17,18^ Further details on the viral genome sequencing and multiplex RT-qPCR variant screening throughout the SARS-CoV-2 waves in Qatar can be found in previous publications.^1,2,4,8,12-16,19-23^

#### Section S3. COVID-19 severity, criticality, and fatality classification

Classification of Coronavirus Disease 2019 (COVID-19) case severity (acute-care hospitalizations),^24^ criticality (intensive-care-unit hospitalizations),^24^ and fatality^25^ followed World Health Organization (WHO) guidelines. Assessments were made by trained medical personnel independent of study investigators and using individual chart reviews, as part of a national protocol applied to every hospitalized COVID-19 patient. Each hospitalized COVID-19 patient underwent an infection severity assessment every three days until discharge or death.

##### Severe COVID-19

Severe COVID-19 disease was defined per WHO classification as a SARS-CoV-2 infected person with “oxygen saturation of <90% on room air, and/or respiratory rate of >30 breaths/minute in adults and children >5 years old (or ≥60 breaths/minute in children <2 months old or ≥50 breaths/minute in children 2-11 months old or ≥40 breaths/minute in children 1–5 years old), and/or signs of severe respiratory distress (accessory muscle use and inability to complete full sentences, and, in children, very severe chest wall indrawing, grunting, central cyanosis, or presence of any other general danger signs)”.^24^ Detailed WHO criteria for classifying Severe acute respiratory syndrome coronavirus 2 (SARS-CoV-2) infection severity can be found in the WHO technical report.^24^

##### Critical COVID-19

Critical COVID-19 disease was defined per WHO classification as a SARS-CoV-2 infected person with “acute respiratory distress syndrome, sepsis, septic shock, or other conditions that would normally require the provision of life sustaining therapies such as mechanical ventilation (invasive or non-invasive) or vasopressor therapy”.^24^ Detailed WHO criteria for classifying SARS-CoV-2 infection criticality can be found in the WHO technical report.^24^

##### Fatal COVID-19

COVID-19 death was defined per WHO classification as “a death resulting from a clinically compatible illness, in a probable or confirmed COVID-19 case, unless there is a clear alternative cause of death that cannot be related to COVID-19 disease (e.g. trauma). There should be no period of complete recovery from COVID-19 between illness and death. A death due to COVID- 19 may not be attributed to another disease (e.g. cancer) and should be counted independently of preexisting conditions that are suspected of triggering a severe course of COVID-19”. Detailed WHO criteria for classifying COVID-19 death can be found in the WHO technical report.^25^

#### Section S4. Phases of the COVID-19 pandemic

The pandemic was categorized into distinct phases based on the level of SARS-CoV-2 incidence and the predominant variant. These phases included the ancestral virus wave (February 28, 2020 - July 31, 2020),^7^ a prolonged low incidence phase with the ancestral virus (August 1, 2020 - January 17, 2021),^2,26^ the alpha wave (January 18, 2021 - March 7, 2021),^27^ the beta wave (March 8, 2021 - May 31, 2021),^28^ a prolonged low incidence delta phase (June 1, 2021 - December 18, 2021),^19,29^ the first (BA.1 & BA.2) omicron wave (December 19, 2021 - February 28, 2022),^6^ the omicron BA.4 & BA.5 wave (March 1, 2022 - September 9, 2022),^22^ and the omicron BA.2.75 & XBB waves (September 10, 2022 - April 21, 2023).^3^

#### Section S5. Comorbidity classification

Comorbidities were ascertained and classified based on the ICD-10 codes as recorded in the electronic health record encounters of each individual in the Cerner-system national database that includes all citizens and residents registered in the national and universal public healthcare system. The public healthcare system provides healthcare to the entire resident population of Qatar free of charge or at heavily subsidized costs, including prescription drugs.

All encounters for each individual were analyzed to determine the comorbidity classification for that individual, as part of a recent national analysis to assess healthcare needs and resource allocation. The Cerner-system national database includes encounters starting from 2013, after this system was launched in Qatar. As long as each individual had at least one encounter with a specific comorbidity diagnosis since 2013, this person was classified with this comorbidity.

Individuals who have comorbidities but never sought care in the public healthcare system, or seek care exclusively in private healthcare facilities, were classified as individuals with no comorbidity due to absence of recorded encounters for them.

**Table S1.**
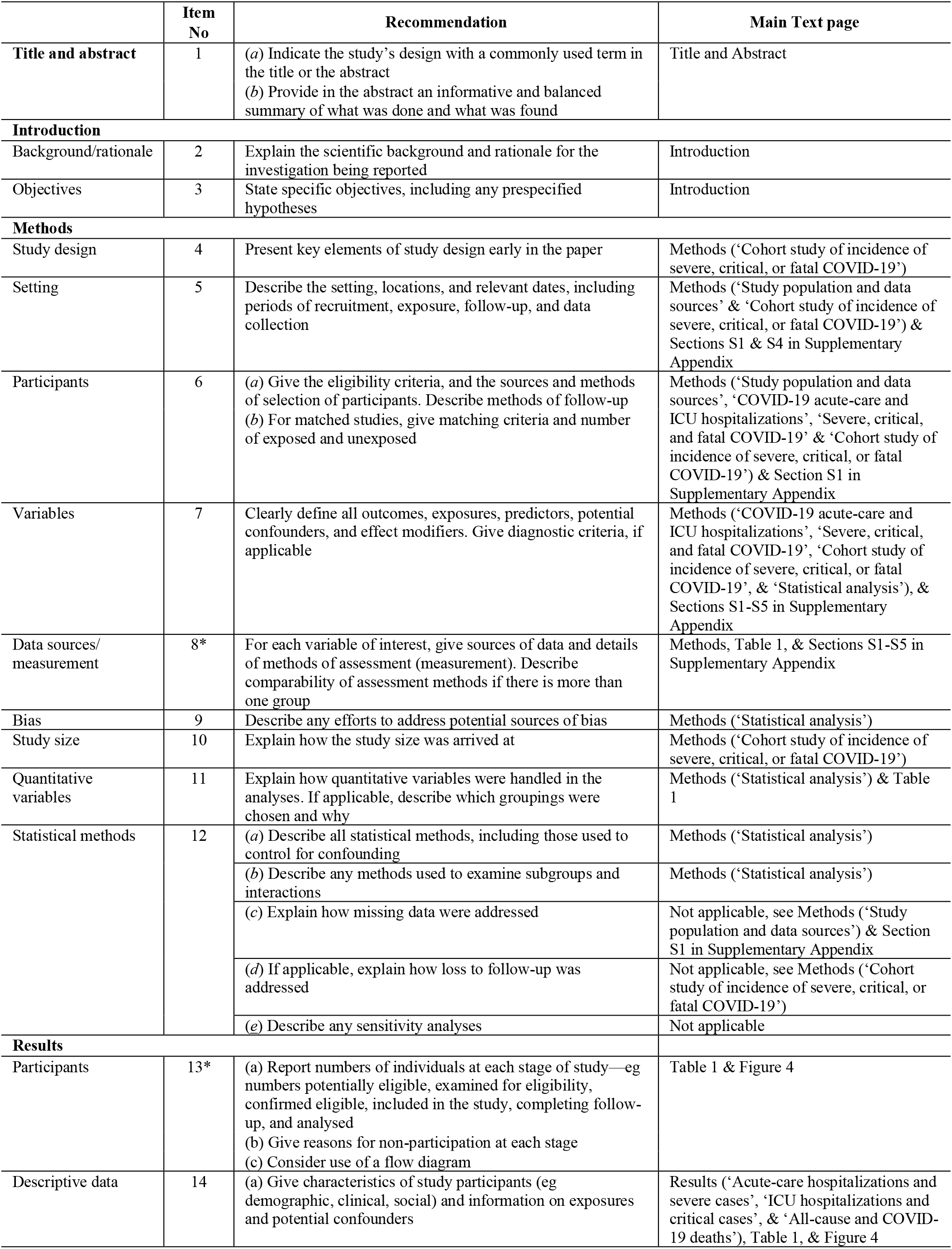
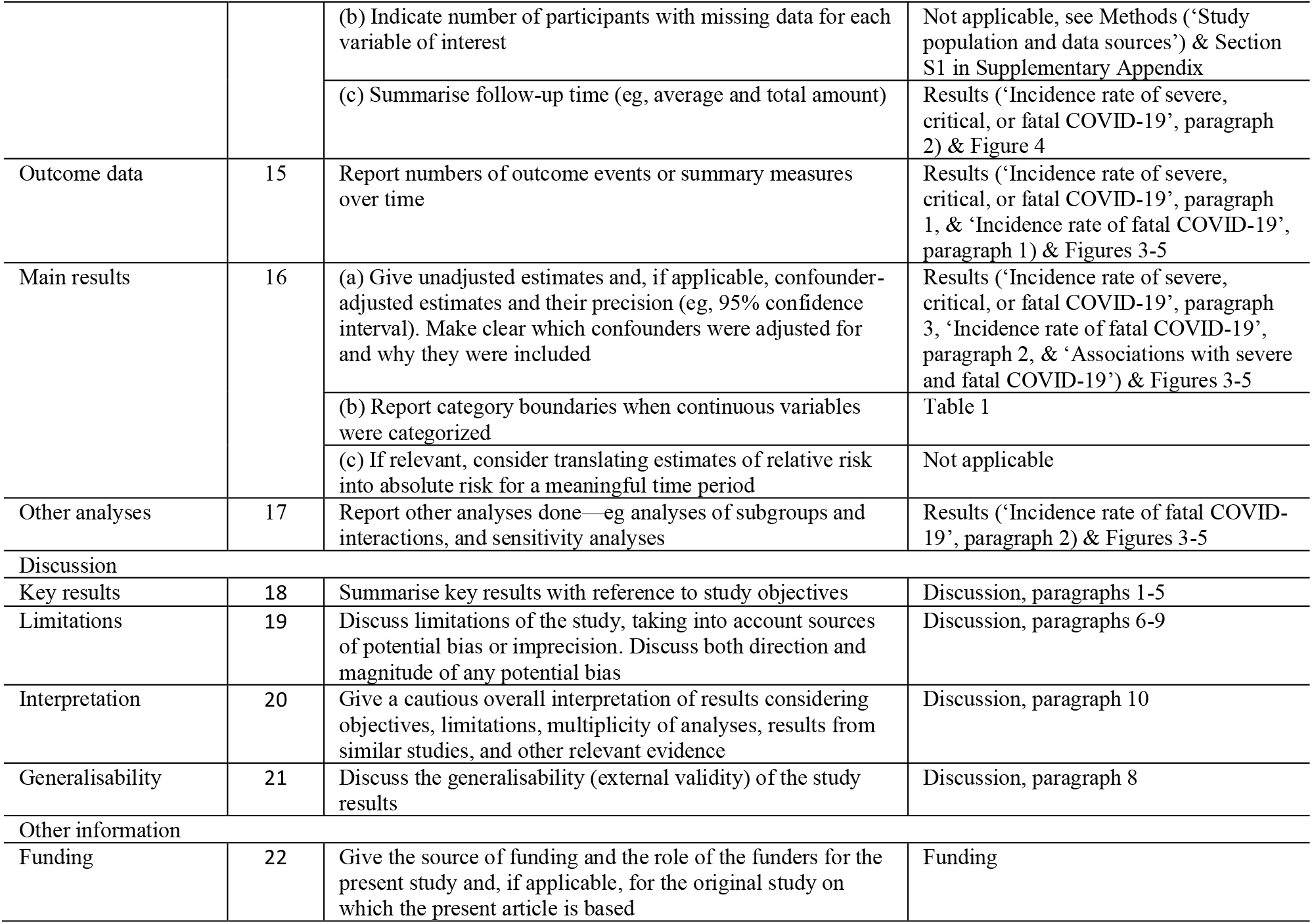
STROBE checklist for cohort studies.

